# Recovery from post-COVID-19 condition and associated factors: the PRIME post-COVID study

**DOI:** 10.1101/2025.10.28.25338941

**Authors:** Demi ME Pagen-Arets, Céline JA van Bilsen, Senne MCE Wijnen, Casper DJ den Heijer, Christian JPA Hoebe, Nicole HTM Dukers-Muijrers

## Abstract

**Background:** Despite the high number of post-COVID-19 condition (PCC) cases worldwide, little is known about recovery and its associated factors yet. This study aimed to estimate the proportion of PCC patients that recover, and identity environment and individual factors.

**Methods:** Longitudinal data of the PRIME cohort were analysed in patients with past infection who felt unrecovered at baseline (November 2021), grouped based on capacity of daily functioning (i.e., moderate to severe problems (PCC-impairedDA) or no or slight problems (PCC-affectedDA)). Proportions of recovery or deterioration at follow-up (after 9 months) were calculated. A range of individual, interpersonal, social network, and social and physical environment factors were assessed for lower likelihood for recovery, using multivariable logistic regression.

**Findings:** 879 patients were analysed. Of patients with PCC-affectedDA (n=602), 222 (36.9%) recovered and 59 (9.8%) deteriorated to PCC-impairedDA. Of patients with PCC-impairedDA (n=277), 30 (10.8%) recovered and 88 (31.8%) improved in daily functioning; recovery was 2.1% when acute illness was >12 months before baseline, and was 12.5% and 13.6% when acute illness was 6-12 and 3-6 months before baseline, respectively. In both PCC groups, individual and environment factors that lowered recovery likelihood included worse physical health (mild/severe fatigue, severe dyspnea, severe symptoms of amnesia, concentration difficulties, muscle pain or -weakness, loss/change of smell/taste), worse mobility (hours spend lying down, problems with walking), worse mental health (depression), worse acute illness (more symptoms, hospitalization/oxygen use), former smoking, living in a rural area, having a relationship, and having more practical social network supporters (in patients with chronic co-morbidities).

**Interpretation:** Only 11%-37% of PCC patients recovered after 9 months, leaving significant room for improvement. Future research should identify modifiable factors and treatments to help assist the recovery of PCC patients.

**Funding:** This study was funded by the Dutch National Institute for Health and Environment, Ministry of Health, Welfare and Sport (Grant numbers: 3910090442/3910105642/3910121041).

## Introduction

At least one out of six people experience persisting symptoms after a SARS-CoV-2 infection (1). These long-term symptoms, referred to as post-COVID-19 condition (PCC), manifests in a wide variety and combinations, of which fatigue, post-exertional malaise and cognitive dysfunction are the most pronounced symptoms (2, 3). The global number of people having PCC is unknown, but estimated at 1.9 million people in the UK (4) and 7.5% (∼25 million) of all US adults (5). For the Netherlands, an estimated 90,000 Dutch adults are living with severely limiting PCC (6).

Patients with PCC deal with its multifaceted impact, which may include limitations in their capacity to perform activities of daily living (7). There is no scientific or practice consensus on how to classify the severity of PCC, but efforts included to account for severity of symptoms (8), a combination between symptoms and quality of life (9), and type of experienced symptoms (10).

In recent years, a growing body of research focused on identifying risk factors for developing PCC, offering insight into subgroups of people who more often develop PCC. These include women, people with more severe acute illness, and people with chronic diseases (11–16). In contrast, there is limited evidence on the proportion of patients that recover and on factors that assist recovery. A recent cross-sectional study estimated that 48.5% of US adults with PCC had recovered (not having these symptoms now) (17), but the duration of PCC was not assessed. Another study showed that 48% of German adults that reported symptoms at 4-12 weeks after acute illness, recovered after 12 weeks (no symptoms) (18). Factors associated with being recovered were younger age (being 40 years or younger), male, employed, mild/moderate symptoms at acute illness (17), being infected with Omicron variant, and not requiring medical care at acute illness (18).

More insight into the progression in terms of recovery or deterioration over time is needed to inform treatment strategies. At current, knowledge is lacking, but it is evident that PCC is a multisystemic and complex illness and recovery from PCC is not universal.

This study aimed to estimate the proportion of patients with PCC – moderate to severe problems with performing daily activities (PCC-impairedDA) or with no or slight problems with performing daily activities (PCC-affectedDA) – who recover or deteriorate over time, and to identify associated factors from the environment to the individual level.

## Method

### Study design and participants

Longitudinal data of the Prevalence Risk factors and Impact evaluation (PRIME) post-COVID study was used (19). In short, PRIME is an observational prospective cohort study, started in 2021, that consists of multiple continued questionnaire rounds. The study assesses various health conditions in relation to COVID-19 illness and PCC. All adults tested for SARS-CoV-2 with a valid test result and email address (recorded in the public health registry) were invited for participation in an online questionnaire. Research platform Crowdtech (ISO 27001 certified) was used for online data collection. Baseline and follow-up data were used for the current analyses. At baseline (November 2021), 12,453 adults participated, whom were invited to participate in the first follow-up questionnaire (August 2022).

Adults with a history of acute COVID-19 illness, meaning a positive SARS-CoV-2 PCR test that was ≥3 months before baseline, who did not feel recovered, and participated at follow-up were selected for this study.

### Individual, interpersonal, social network, and social and physical environment factors

The baseline questionnaire covered individual (demographics, general physical health, symptoms at baseline, mobility, (severity of) acute COVID-19 illness, vaccination status, mental and social health, lifestyle), interpersonal (steady partner, home situation, social network), and societal (physical environment) factors (table 1), which were all explored for their association with recovery from PCC. The questionnaire did not cover duration of PCC. We considered time since first acute COVID-19 illness as a suitable proxy for potential duration of PCC, as the risk of new-onset PCC has been suggested to be lower following second infection compared to first (20).

**Table 1.** Overview of all factors tested to be associated with changing post-COVID-19 condition (recovered, PCC-affectedDA, PCC-impairedDA) status from baseline to follow-up.

| Individual level | Measured at baseline |  |
| --- | --- | --- |
|  | Currently | Retrospectively |
| *Demographic factors* |  |  |
| Sex (men/women) | x |  |
| Age (years) | x |  |
| Born in the Netherlands (yes/no) | x |  |
| Level of education (practically/theoretically trained) | x |  |
| Paid job (yes/no) | x |  |
| *General (physical) health* |  |  |
| Obese (body mass index (BMI) $\geq 30$ kg/m<sup>2</sup>; yes/no) | x | |
| Diabetes (yes/no) |  | x |
| Chronic obstructive pulmonary disease (COPD; yes/no) |  | x |
| Asthma (yes/no) |  | x |
| Lung disease (yes/no) |  | x |
| Heart disease (yes/no) |  | x |
| General health score (EuroQol-visual analogue scales (EQvas; range 0-100) |  |  |
| Year before test |  | x |
| At baseline | x |  |
| Fatigue (checklist individual strength-subscale subjective fatigue (CIS-fatigue) (21); normal/mild/severe) | x |  |
| Dyspnea (modified medical research council (mMRC) (22); normal to mild/severe) | x |  |
| *Symptoms at baseline* |  |  |
| Amnesia (no/severity 1-5/severity $\geq 5$-10) | x | |
| Brain fog (no/severity 1-5/severity $\geq 5$-10) | x | |
| Concentration difficulties (no/severity 1-5/severity ≥5-10) | x |  |
| Confusion (no/severity 1-5/severity ≥5-10) | x |  |
| Dizziness (no/severity 1-5/severity ≥5-10) | x |  |
| Headache (no/severity 1-5/severity ≥5-10) | x |  |
| Palpitations (no/severity 1-5/severity ≥5-10) | x |  |
| Increased resting heart rate (no/severity 1-5/severity ≥5-10) | x |  |
| Joint pain (no/severity 1-5/severity ≥5-10) | x |  |
| Muscle pain or -weakness (no/severity 1-5/severity ≥5-10) | x |  |
| Sleeping problems (no/severity 1-5/severity ≥5-10) | x |  |
| Loss/change of smell/taste (no/severity 1-5/severity ≥5-10) | x |  |
| *Mobility* |  |  |
| Time spend lying down (hours/day) | x |  |
| Problems with walking (item EuroQol 5-Dimensions (EQ5D) (23); no to slight/moderate to (very) severe) | x |  |
| *(severity of) acute COVID-19 illness* |  |  |
| Symptoms when tested (total number from 0-44) |  | x |
| Hospitalization/oxygen use at home or hospital (yes/no) |  | x |
| Months since SARS-CoV-2 test (based on date of positive test; 3-6 months/6-12 months/≥12 months) | x |  |
| *Vaccination status* |  |  |
| COVID-19 vaccinated (yes/no) | x |  |
| *Mental and social health* |  |  |
| Depression (patient health questionnaire-9 (PHQ-9) (24); yes/no) | x |  |
| Loneliness (De Jong-Gierveld (25); not lonely/moderately lonely/severely lonely) | x |  |
| Emotional loneliness (yes/no) | x |  |
| Social loneliness (yes/no) | x |  |
| Anxiety (yes/no) | x |  |
| *Lifestyle* |  |  |
| Smoking behaviour (never/former/current) | x |  |
| **Interpersonal level** |  |  |
| *Steady partner* |  |  |
| Having a relationship (yes/no) | x |  |
| *Home situation* |  |  |
| Living alone (yes/no) | x |  |
| Children (yes/no) | x |  |
| *Social network factors (name generator method) (26)* |  |  |
| Size of social network (number of relations from 0-45) | x |  |
| Number of members giving emotional support | x |  |
| Number of members giving practical support | x |  |
| Number of members giving informational support | x |  |
| Network diversity (no family, but friends or acquaintances or caregivers/only family, no friends or acquaintances or caregivers/family and friends, no acquaintances or caregivers/family and friends and acquaintances or caregivers/family and acquaintances or caregivers, no friends) | x |  |
| Network density (best friends know family (yes/no)) | x |  |
| **Societal level** |  |  |
| *Physical environment* |  |  |
| Urbanity of living area (based on postal code; (very) strongly urban/moderately urban/little urban/rural) | x |  |
| PCC-affectedDA=post-COVID-19 condition with no or slight problems with performing daily activities; PCC-impairedDA=post-COVID-19 condition with moderate to severe problems or unable to perform daily activities. |  |  |

### Outcome measure

The outcome measure was the PCC status at follow-up, based on a) whether participants currently felt fully recovered from their COVID-19 infection and b) their current capacity of daily functioning. The latter was taken to reflect severity of PCC and was based on the EuroQol-5 Dimensions (EQ5D) item (on the domain usual activities) regarding the health today. The answers to this item (no, slight, moderate, severe problems, or not able to perform daily activities) were dichotomized into no or slight problems (in short: PCC-affectedDA) and moderate to severe problems or not able to perform daily activities (in short: PCC-impairedDA). At baseline and follow-up, patients were grouped as PCC-affectedDA or PCC-impairedDA, or recovered (at follow-up only).

### Statistical analysis

Proportions and 95% confidence intervals (CI) of participants with no PCC, PCC-affectedDA and PCC-impairedDA at follow-up were calculated, also stratified for time since first acute COVID-19 illness, grouped into 3-6 months, 6-12 months and ≥12 months.

Chi-square or Fisher’s Exact tests (for categorical) and analysis of variance (ANOVA; for continuous) variables were used to assess differences in the prevalence of factors by PCC status.

Multivariable logistic regression analyses were performed to assess the factors (measured at baseline) for their association with PCC status at follow-up (9 months later), adjusted for confounders and stratified by baseline PCC status. PCC status could thus either improve to recovered or better daily functioning, or could remain unaltered over time. Unaltered status was selected as the reference group. Included confounders were age (years), time since testing (3-6 months, 6-12 months, and ≥12 months) and having had another COVID-19 infection between baseline and follow-up (yes/no).

Interactions between statistically significant factors and sex (men/women) and having chronic conditions (yes/no) were tested to evaluate possible heterogeneity of effects of the identified factors. Analyses were performed using Statistical Package for Social Sciences (SPSS version 27, IBM, Armonk, USA); a p-value <0.05 was considered statistically significant.

### Ethical statement

This study was waived by the Medical Ethical Committee of Maastricht University Medical Centre+ (METC2021-2884), as the Medical Research Involving Human Subjects Act (WMO) did not apply. This study was registered at ClinicalTrials.gov Protocol Registration and Results System (NCT05128695).

### Data availability

The data that support the findings of this study are available on request from the head of the data-archiving South Limburg Public Health Service for researchers who meet the criteria for access to confidential data. Requests to access the datasets should be directed to. The data are not publicly available because the data contains potentially identifying patient information.

## Results

Overall, 3,922 test positive participants in the cohort had complete data at both baseline and follow-up. Participants were excluded from further analysis when they felt recovered at baseline (n=1,607), tested SARS-CoV-2 negative until the follow-up questionnaire (n=621), or tested SARS-CoV-2 positive <3 months prior to completing the baseline or follow-up questionnaire (n=122). Also, participants that were likely not the intended invitee (i.e., mismatch on sex or test result between baseline and follow-up; n=156) were excluded. The final study population in analyses consisted of 879 participants, of whom 602 (68.5%) had PCC-affectedDA and 277 (31.5%) had PCC-impairedDA at baseline.

### Baseline characteristics

People with PCC-affectedDA differed from people with PCC-impairedDA on diverse aspects: generally PCC-impairedDA scored worse on having a paid job, chronic conditions (obesity, COPD, asthma, heart disease), general health before their COVID-19 test and at baseline, current fatigue, dyspnea, mobility (hours spend lying down; problems with walking), depression, emotional loneliness, severity of acute illness (more symptoms, hospitalization or oxygen use), current symptoms (amnesia, brain fog, concentration difficulties, confusion, dizziness, headache, palpitations, increased resting heart rate, joint pain, muscle pain or -weakness, sleeping problems, and loss/change of smell/taste), having more social (emotional, practical, informational) network supporters (table 2).

**Table 2.**
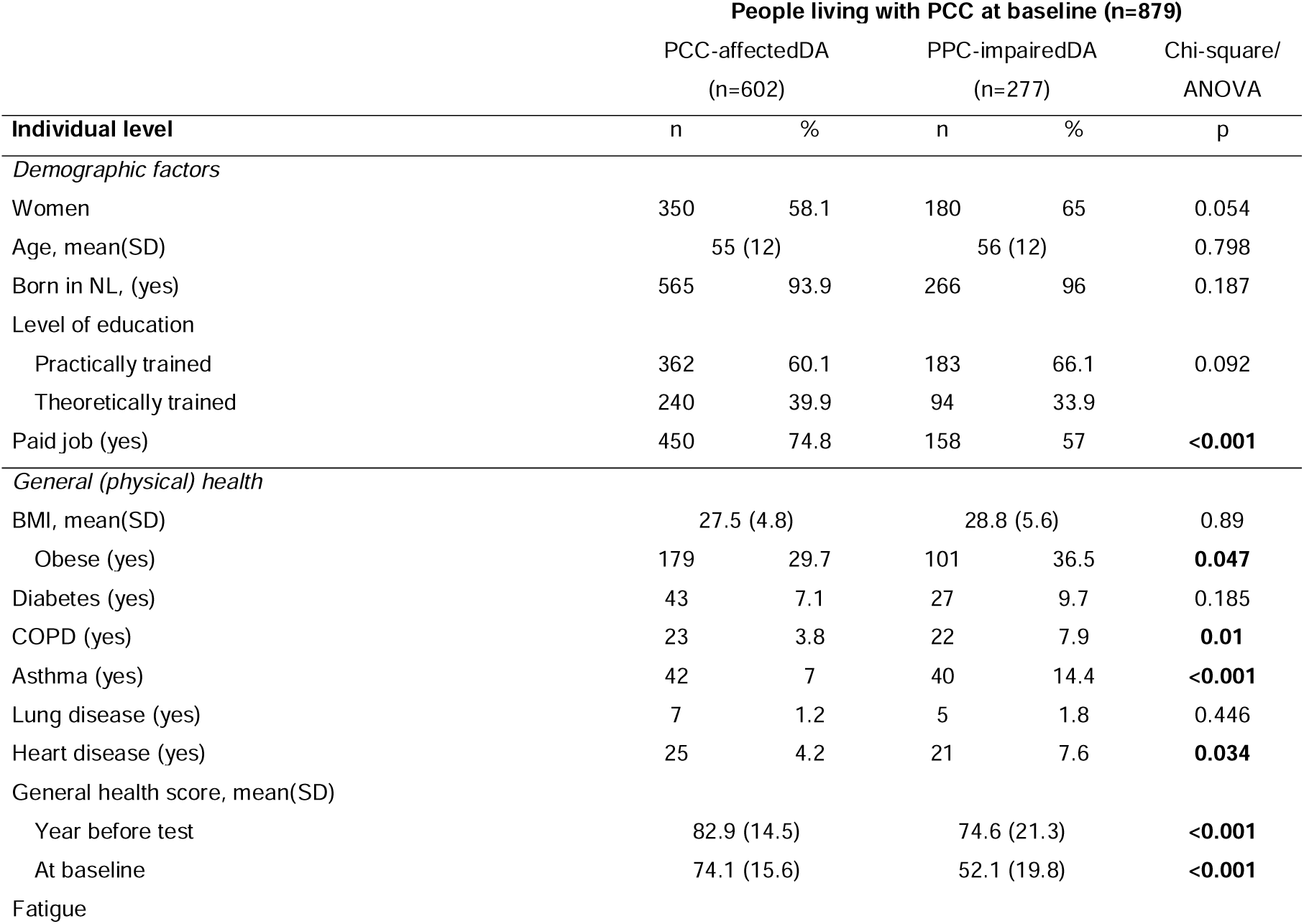
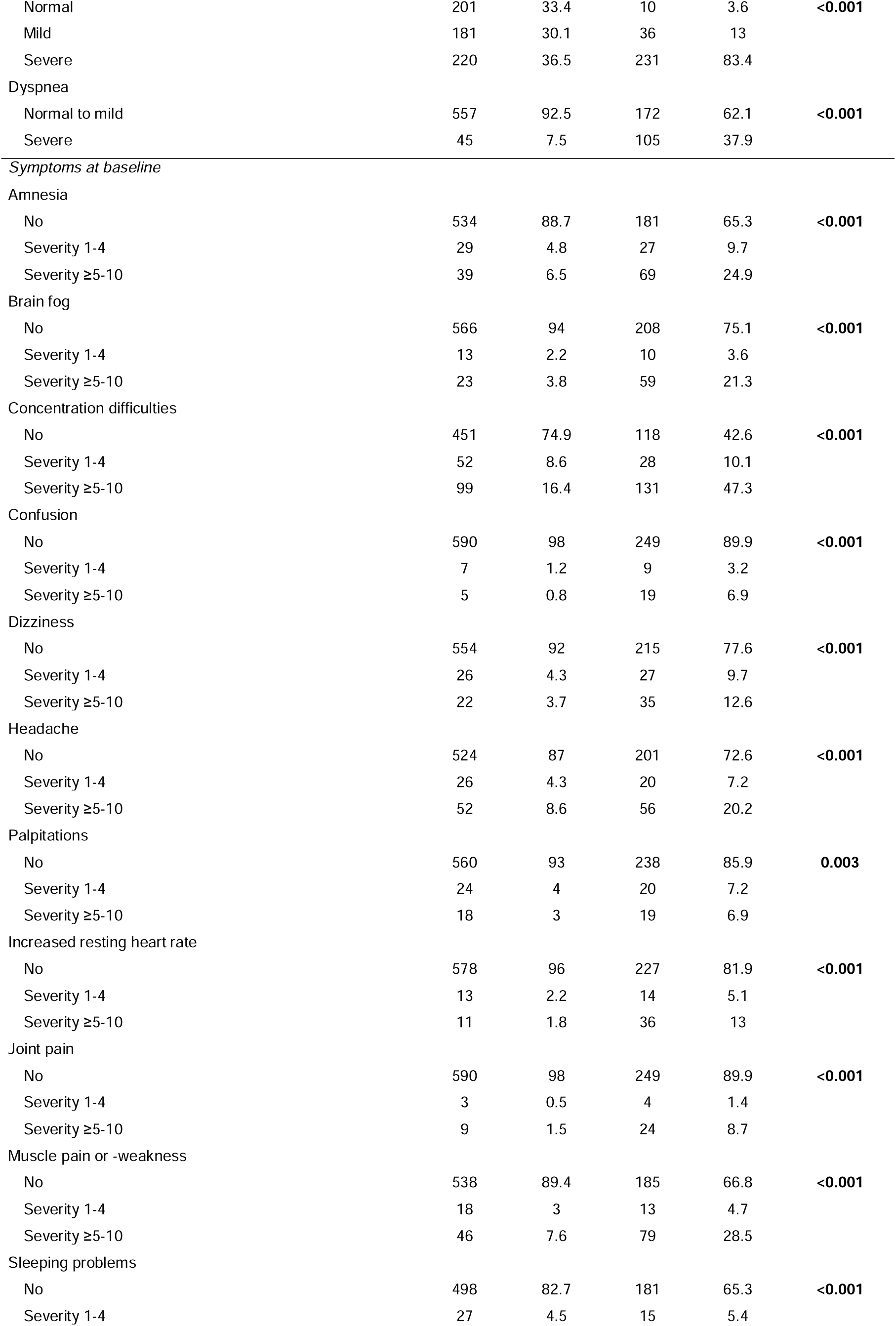
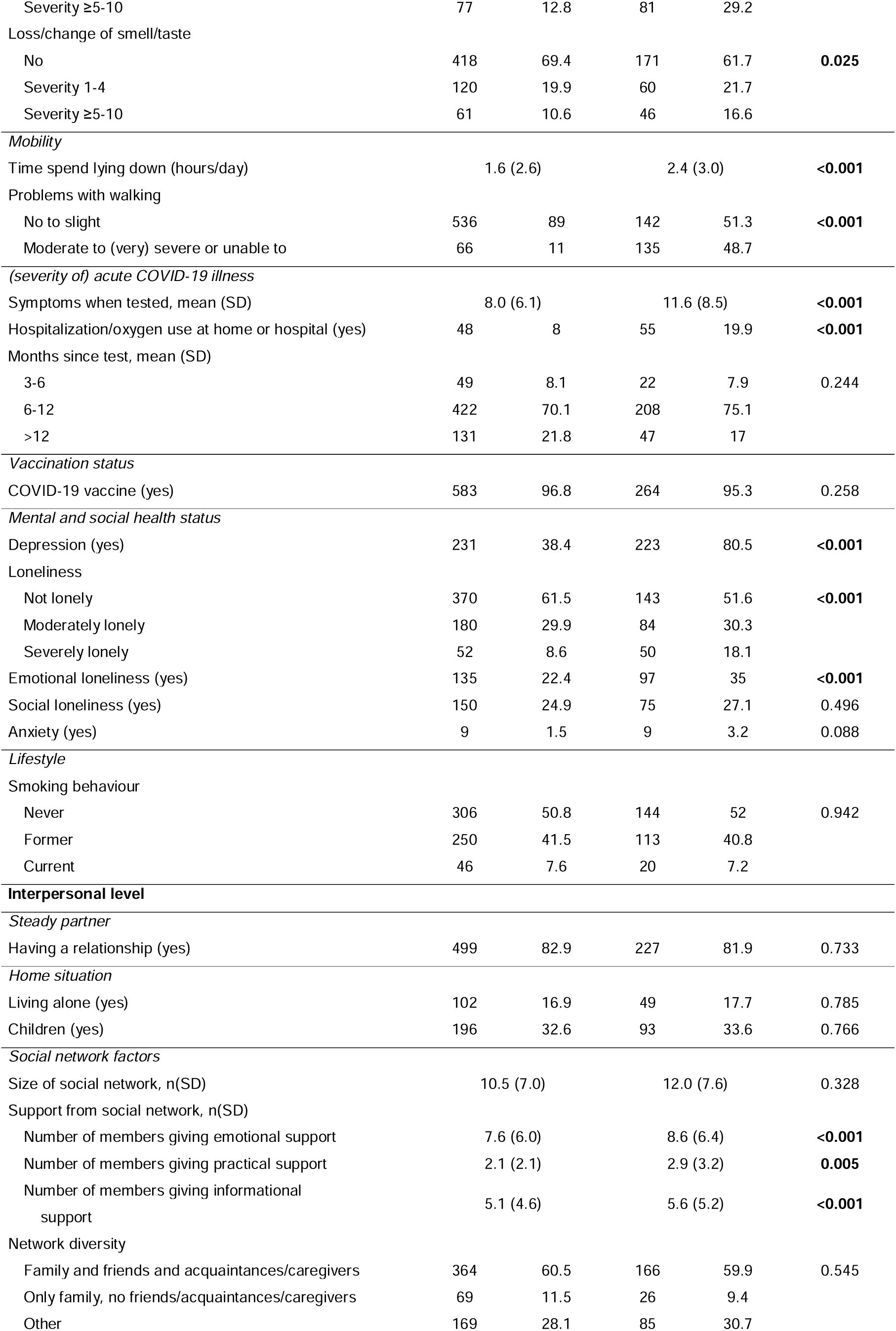
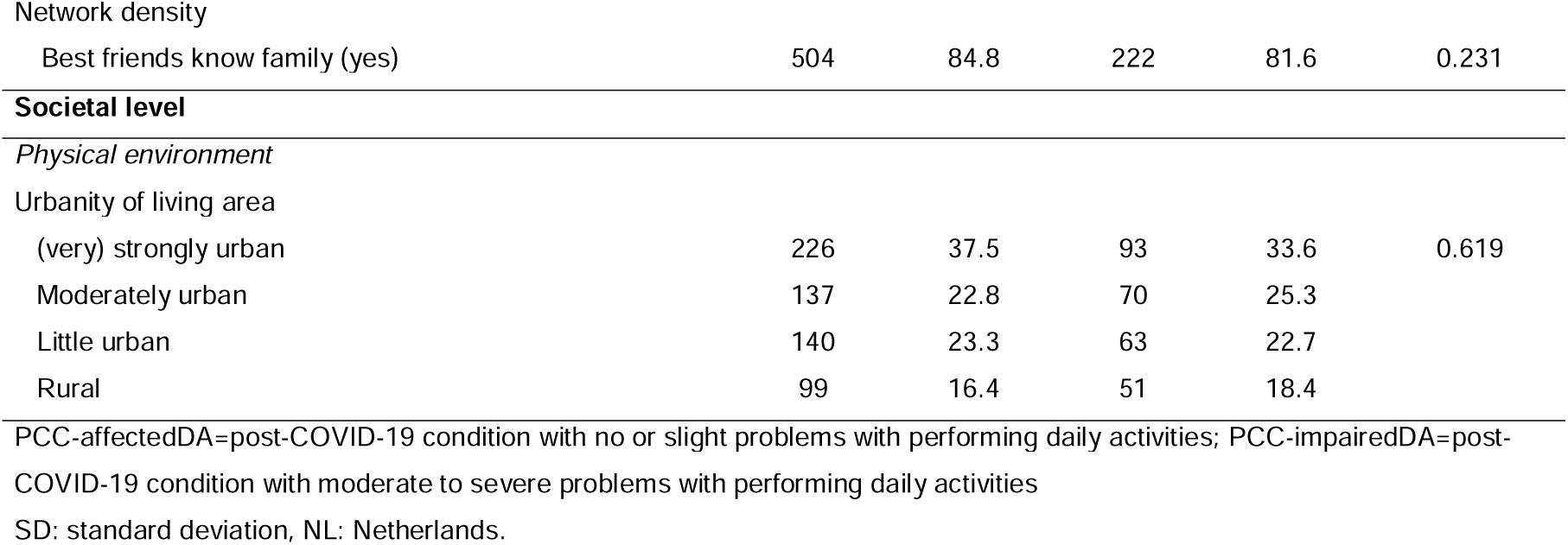
Baseline characteristics of people with PCC-affectedDA and PCC-impairedDA.

### Proportion of patients who changed to different PCC groups at follow-up

Of patients with PCC-affectedDA at baseline (n=602), 222 (36.9% [95% CI:30.0%-40.7%]) recovered, 321 (53.3% [95% CI:49.3%-57.3%]) persisted having PCC-affectedDA, and 59 (9.8% [95% CI:7.4%-12.2%]) deteriorated to PCC-impairedDA at follow-up (figure 1). The proportion of recovery did not differ by duration of PCC (Chi-square p=0.166); for those with first acute COVID-19 illness 3-6 months before baseline it was 44.9% [95% CI:40.9%-48.9%], 6-12 months 36.0% [95% CI:32.2%-39.9%], and ≥12 months 36.6% [95% CI:32.8%-40.5%].

**Figure 1.**
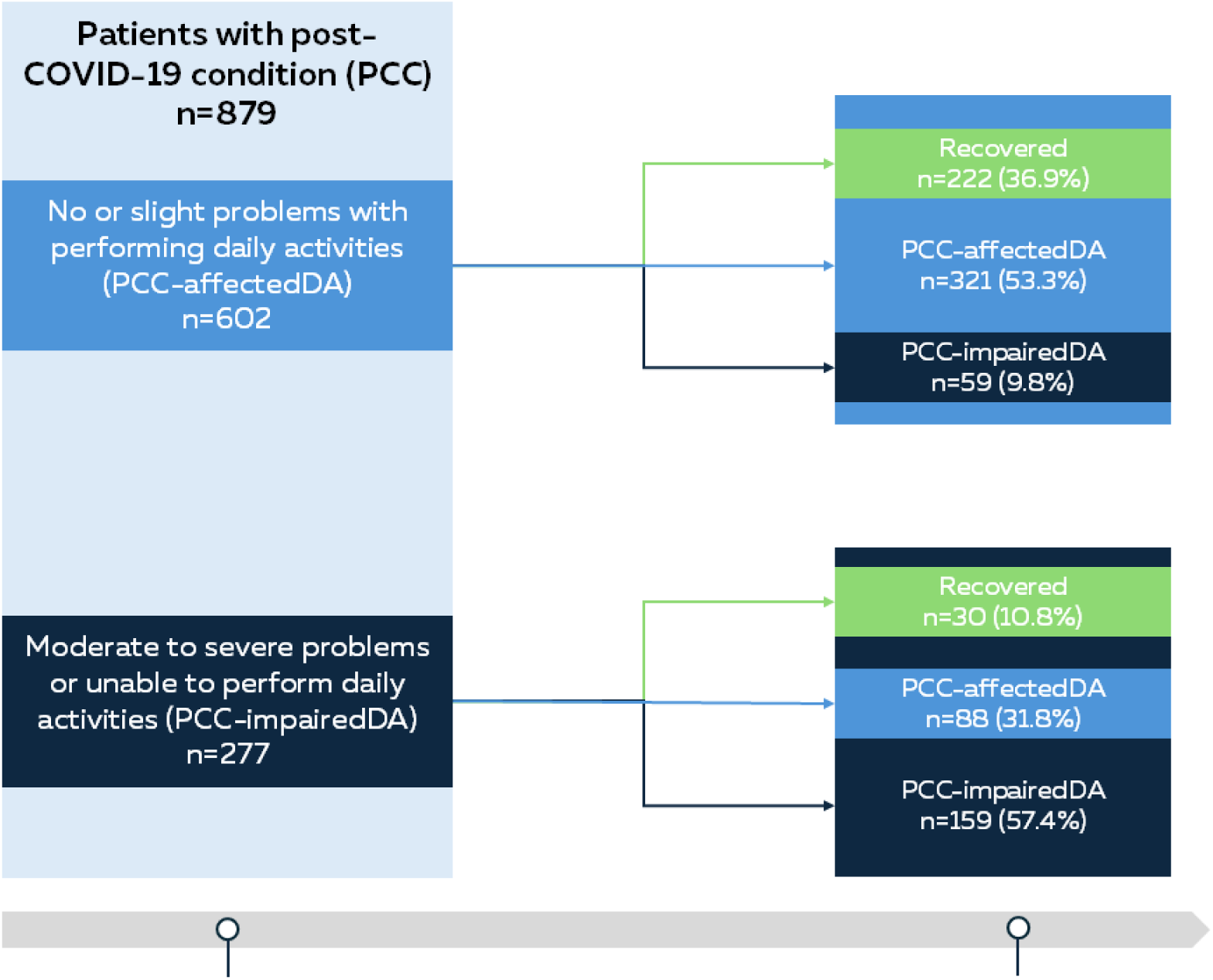
Overview of the shift in post-COVID-19 condition (PCC) status from baseline to follow-up.

Of patients with PCC-impairedDA at baseline (n=277), 30 (10.8% [95% CI:7.2%-14.5%]) recovered, 88 (31.8% [95% CI:26.3%-37.3%]) improved somewhat to PCC-affectedDA, and 159 (57.4% [95% CI:51.6%-63.2%]) persisted having PCC-impairedDA at follow-up (figure 1). Recovery was 2.1% [95% CI:0.4%-3.8%] when first acute COVID-19 illness was long ago (≥12 months before baseline), and was 12.5% [95% CI:8.6%-16.4%] (p=0.036) and 13.6% [95% CI:9.6%-17.7%] (p=0.092) when acute illness was 6-12 and 3-6 months before baseline, respectively.

### Factors associated with recovery

A detailed description of the proportions of all factors within each PCC group (i.e., recovered, PCC-affectedDA and PCC-impairedDA) at baseline and follow-up is presented in supplementary tables 1 and 2.

#### In patients with PCC-affectedDA

On individual level, the following health factors were associated with a lower likelihood for recovery: worse general health, worse physical health (mild/severe fatigue, severe symptoms of amnesia, concentration difficulties, muscle pain or -weakness, loss/change of smell/taste) worse mobility (hours spend lying down), worse acute illness (more symptoms when tested, hospitalization/oxygen use), worse mental health (depression), and former smoking. Having a relationship and having more practical social network supporters (in patients with chronic co-morbidities) on interpersonal level, and living in a rural area on societal level were associated with a lower likelihood for recovery (figure 2).

**Figure 2.**
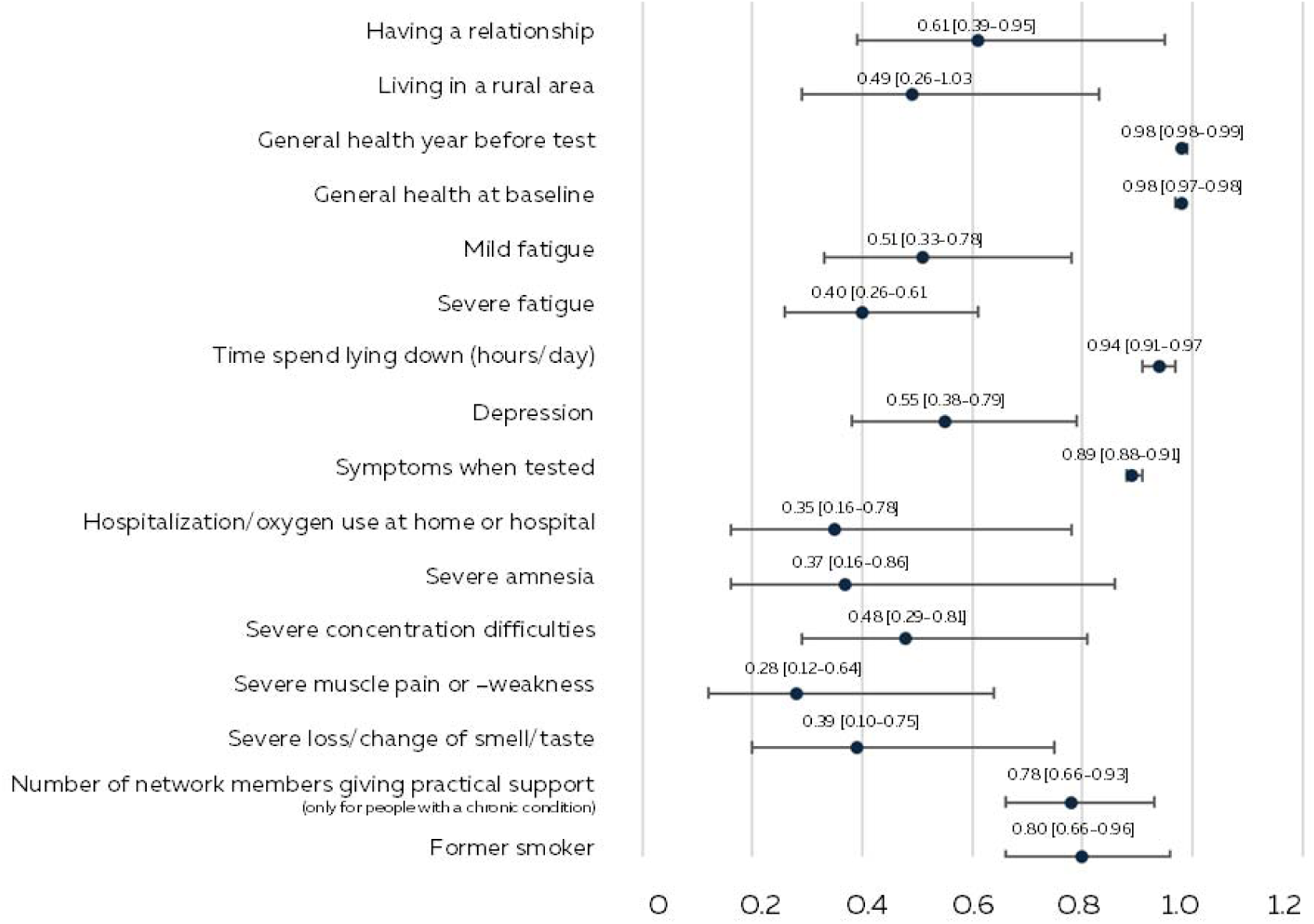
Forest plot of factors associated with lower likelihood for recovery from post-COVID-19 condition with no or slight problems with performing daily activities (PCC-affectedDA).

#### In patients with PCC-impairedDA

On individual level, factors associated with a lower likelihood for recovery were: worse general health, worse physical health (mild/severe fatigue, severe dyspnea, severe symptoms of amnesia or concentration difficulties), worse mobility (moderate to (very) severe problems with walking), worse acute illness factors (more symptoms when tested), and worse mental health (depression) (figure 3).

**Figure 3.**
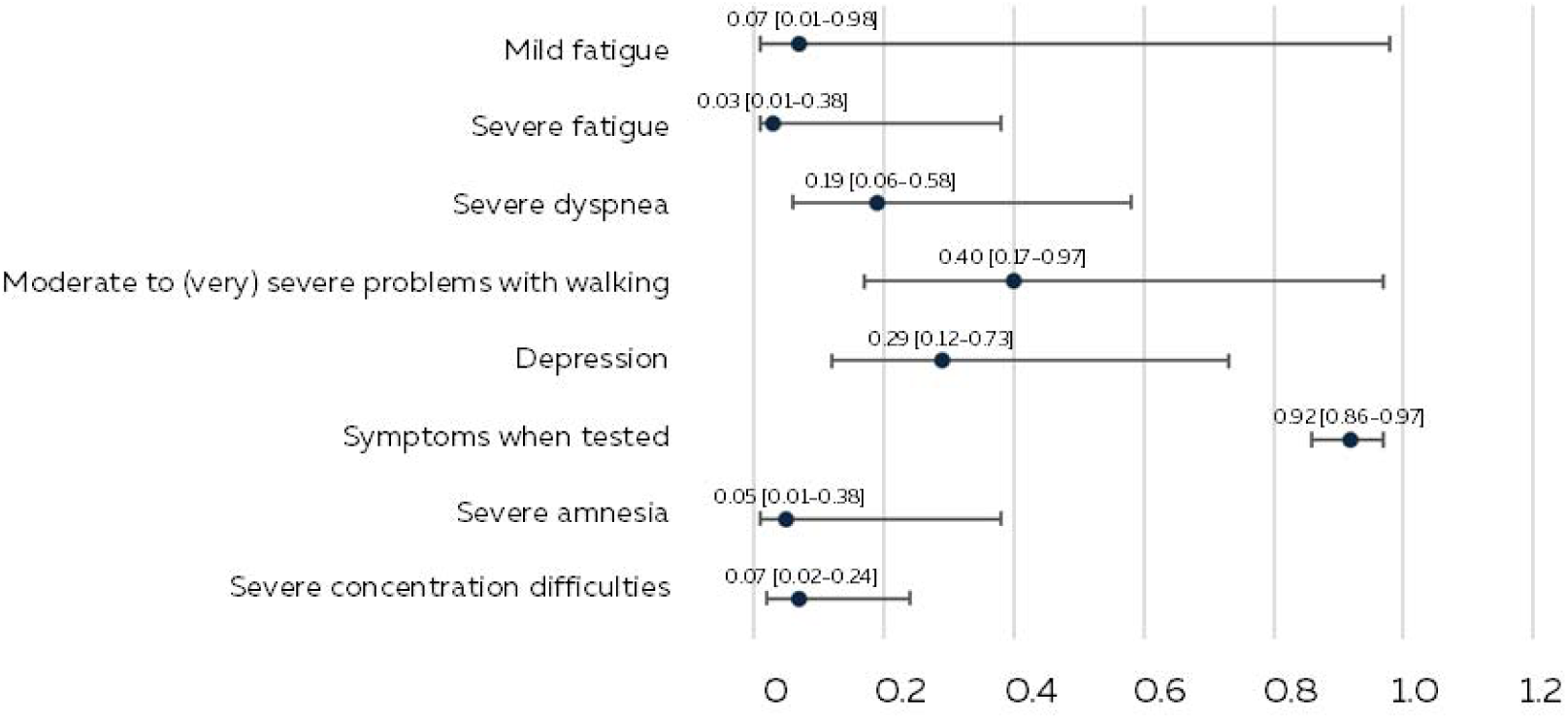
Forest plot of factors associated with lower likelihood for recovery from post-COVID-19 condition with moderate to severe problems with performing daily activities (PCC-impairedDA).

On individual level, improvement to PCC-affectedDA was negatively associated with: worse general health, worse physical health (mild/severe fatigue, symptoms of severe brain fog or concentration difficulties), worse acute illness factors (hospitalization/oxygen use), worse mental health (depression, moderate loneliness or emotional loneliness (in women and patients without chronic co-morbidities)) (figure 4).

**Figure 4.**
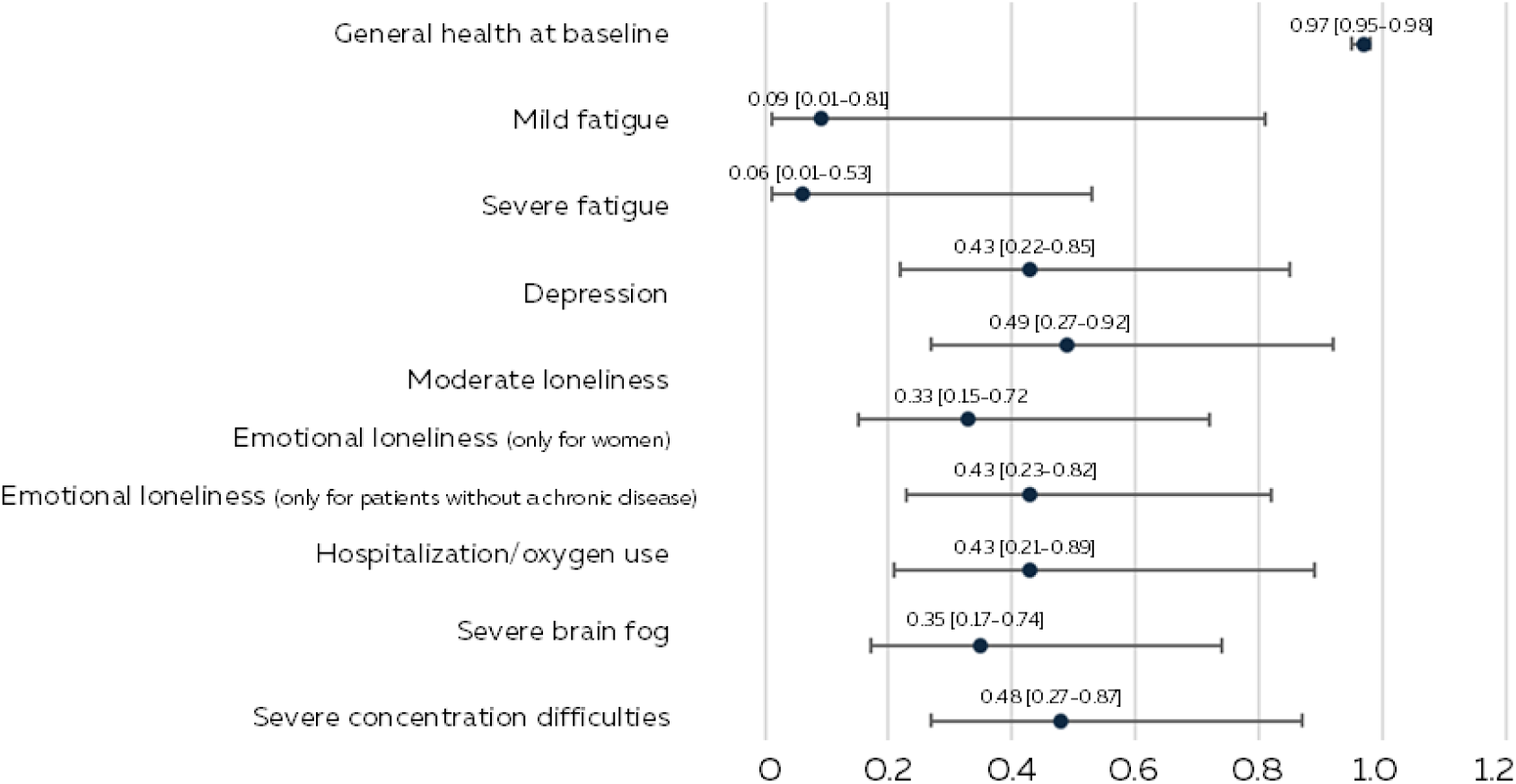
Forest plot of factors associated with a lower likelihood for improvement from post-COVID-19 condition with moderate to severe problems with performing daily activities (PCC-ImpairedDA) to post-COVID-19 condition with no or slight problems with performing daily activities (PCC-affectedDA).

### Factors associated with deterioration in patients with PCC

Factors associated with deterioration to PCC-impairedDA were severe fatigue (adOR=2.68 [95%CI:1.23-5.81]), severe dyspnea (adOR=2.92 [95% CI:1.33-6.42]), and moderate to (very) severe problems with walking (adOR=2.71 [95% CI:1.32-5.54]) (supplementary table 1). A worse general health score was positively associated with deterioration (adOR=1.02 [95%CI: 1.01-1.04]) (supplementary table 1).

### Description of PCC groups at follow-up

Demographic and health characteristics were described for people who recovered, had PCC-affectedDA or had PCC-impairedDA at follow-up (figure 5).

**Figure 5.**
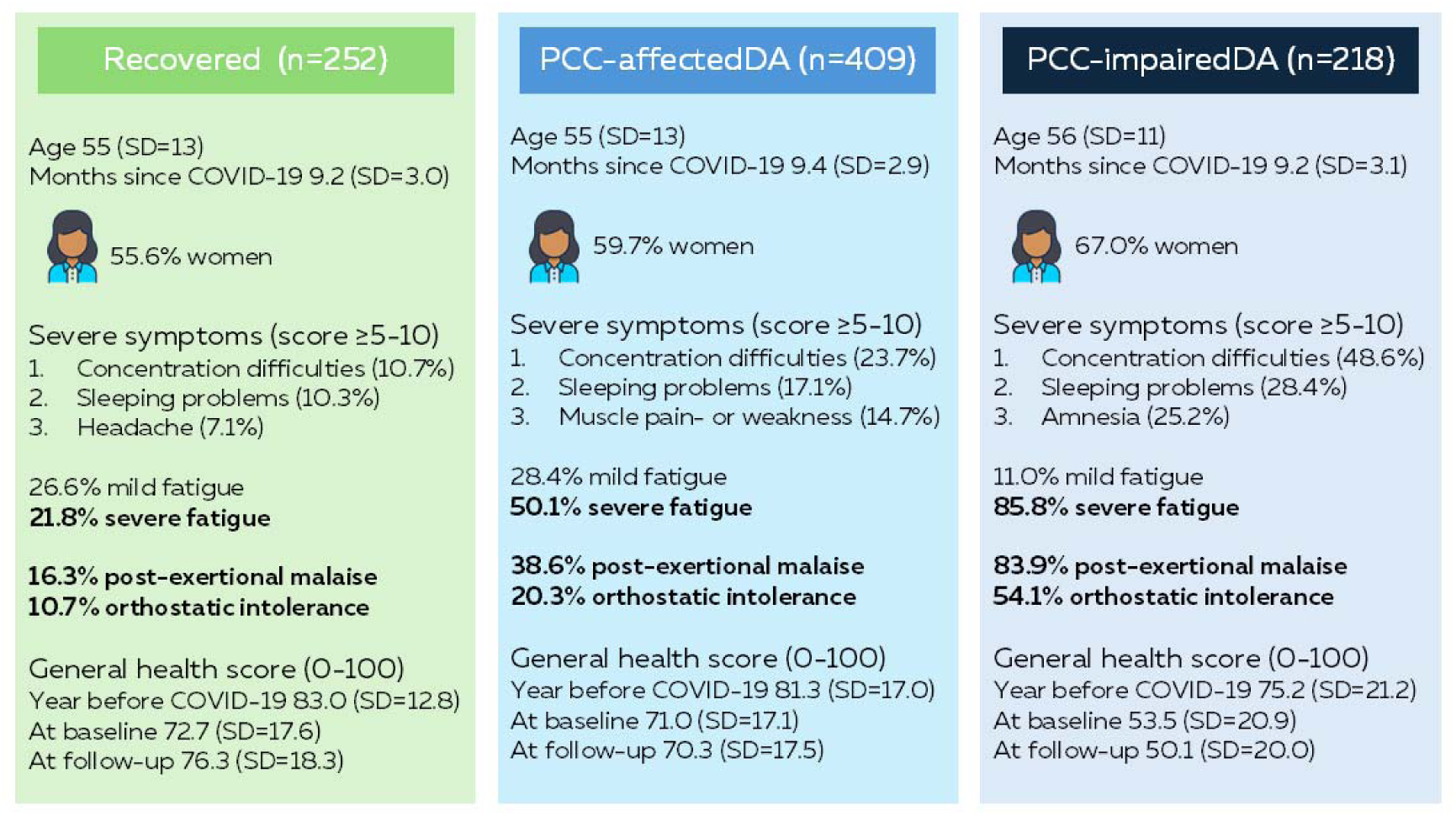
Description of PCC groups (i.e., recovered, post-COVID-19 condition with no or slight problems with performing daily activities (PCC-affectedDA), and post-COVID-19 condition with moderate to severe problems with performing daily activities (PCC-impairedDA)) at follow-up.

Patients with PCC-impairedDA were more often women (67.0%), compared to PCC-affectedDA patients (59.7%) and to recovered people (55.6%; p=0.039). In patients with PCC-impairedDA, 48.6% had post-exertional malaise (PEM), 54.1% had orthostatic intolerance (OI), and 85.8% had severe fatigue, which was higher than in patients with PCC-affectedDA (38.6% PEM; p<0.001, 20.3% OI; p<0.001, and 50.1% severe fatigue; p<0.001) or in people who recovered (16.3% PEM; p<0.001, 10.7% OI; p<0.001, and 21.8% severe fatigue; p<0.001). In all PCC groups, concentration difficulties and sleeping problems were the most reported severe symptoms and these symptoms were most often reported in patients with PCC-impairedDA compared to patients with PCC-affectedDA or people who recovered. The general health score was the highest a year before acute COVID-19 illness, for all PCC groups. For patients who recovered, the general health score indeed improved, while general health on average had deteriorated in those who had PCC-affectedDA or PCC-ImpairedDA at follow-up.

## Discussion

One in three PCC-affectedDA patients and one in ten PCC-impairedDA patients, participating in the PRIME cohort, had recovered in a nine-month period. Identified factors associated with a lower likelihood for recovery included a range of individual and environment factors; worse general, physical and mental health, and worse acute COVID-19 illness.

Recently, the National Academies of Sciences, Engineering, and Medicine (NASEM) definition of PCC was formed, emphasizing that an important feature in defining PCC lies in the possibility that it can range from mild to severe (27). Nevertheless, currently, no proposed definition of PCC includes a minimum level of symptom severity or functional impairment (28). The definition of PCC used in our study, based on the capacity of daily functioning, allowed for differentiation by PCC severity. Notable differences between patients with impaired daily functioning and patients not or slightly affected in daily functioning were observed; including higher occurrence of problems with walking (48.7% in vs. 11.0%), depression (80.5% vs. 38.4%) and presence of severe symptoms (e.g., concentration difficulties 47.3% vs. 16.4%). Thereby, our study suggests a way to define PCC severity by its accompanying impairment of daily living, instead of classifying PCC severity groups based on symptom clusters alone (8–10).

Comparison of our results on recovery proportions with previous studies is nearly impossible, as there is no consensus on a PCC definition nor on recovery. Different use of study designs and inclusion of diverse study populations further complicate comparison of research output. We demonstrated that only 37% of patients with PCC-affectedDA recovered, and only 11% of patients with PCC-impairedDA recovered in 9 months, which is less than suggested by previous studies which estimated recovery to be around 50% (17, 18). One explanation could be the inclusion of more patients with longer PCC duration, since longer time since first COVID-19 illness was negatively associated with recovery in our study. In previous studies, participants were classified as having PCC when they reported having symptoms 4-12 weeks after infection, or the presence of any symptoms lasting 3 months or longer that they did not have prior to having COVID-19 (17, 18). Thus, these PCC definitions were focused on presence of symptoms alone, instead of (our study) the experienced recovery after COVID-19 infection combined with the current degree of impairment of daily living. Furthermore, while other studies defined recovery upon absence of current symptoms, our study demonstrated that people who felt recovered, indeed had improved health, yet still experienced (although less often) severe symptoms or key symptoms as PEM and OI. Possibly, the patients experienced their condition as better than before and the still present symptoms are more or less manageable. On the other hand, patients in our study who were unrecovered, had either (or both) symptoms or impairment in daily functioning. This questions whether only asking for just presence of symptoms in clinical practice is a good indicator to identify patients’ recovery, without some assessment of impact on functioning.

People who felt recovered had their general health score during the course of PCC improved, although, it is notable that their score was still under the Dutch average reference value (score of 86.9, measured in 2016) (29) and lower than the average in people who never reported to have had COVID-19 before (score of 79.2, measured in 2021) (7).

Overall, it must be acknowledged that defining PCC and recovery are complex. Yet, alignment of definitions used in practice is urgently needed to enhance comparability between studies to estimate the proportion of people with PCC that are able to recover over time.

Factors that lowered the likelihood for recovery - or even were associated with deterioration - included individual (worse general health, physical health (mild/severe fatigue, severe dyspnea, severe symptoms of amnesia, brain fog, concentration difficulties, muscle pain or -weakness, loss/change of smell/taste) worse mobility (hours spend lying down, moderate to (very) severe problems with walking), worse acute illness (number of symptoms when tested, hospitalization/oxygen use), worse mental and social health (depression, moderate or emotional loneliness) and former smoking), interpersonal (having a relationship, having more practical social supporters in patients with chronic co-morbidities) and societal (living in a rural area) factors. PCC-impairedDA patients who were more often women or had severe symptoms (i.e., concentration difficulties, sleeping problems, amnesia, fatigue, PEM and OI) were less likely to recover.

We demonstrated that a worse health before and during PCC are associated with a lower likelihood for recovery, which is unfortunately largely unchangeable and requires biological targets to be treated.

Nevertheless, results of the current study highlight the importance of a good overall health, which can be seen as a starting point for prevention. This study was not designed to reveal therapeutic targets for recovery, and we recommend future biomedical and therapeutic research to give insight into possible biological targets to ultimately treat PCC.

### Strengths and limitations

We were able to offer insights into recovery trajectories in a large set of PCC patients, using longitudinal, population-based data to estimate the proportion of PCC patients (having PCC for 3 up to 21 months) who subsequently recover. Additionally, a wide range of individual and environment health factors were evaluated for their association with recovery. The extensive measures used provided detailed information on various mental, physical, and social health conditions, and social and physical living environment factors; most measured with validated scales. To the best of our knowledge, this is the first study that demonstrates distinct PCC groups based on capacity of daily functioning, by combining the outcome of two relatively simple questions, which could be used in future studies and clinical practice.

A limitation is the lack of biomedical data, clinical assessments and biomarkers. Due to the nature of the cohort, availability of data was by self-report. Even so, various measures such as fatigue and impairments of daily living are difficult to determine clinically or with biomedical assessments. Another limitation is loss-to-follow-up in the PRIME study, possibly leading to selection or attrition bias, which might be differential by PCC severity. PCC patients with profound impairment of daily living are possibly less capable to complete the relatively long questionnaire in this study, and therefore we consider it likely that the most severely impacted PCC patients are underrepresented in this study.

This might have resulted in an overestimation of recovery in adults with PCC. Nevertheless, we included a substantial sample of PCC patients with severe impact in daily functioning. The exact duration of PCC is unknown. However, we considered the time since first acute COVID-19 illness as a proxy, demonstrating that PCC patients with severe impact on daily functioning who had acute COVID-19 illness ≥12 months before baseline were least likely to recover. Furthermore, the measure on impairment of daily living reflected the situation ‘’today’’, having its limitations to indicate participants general state, specifically in PCC which is known for its intermittent symptoms; hence recovery might have been underestimated when patients were currently in remission or overestimation when patients were currently having a relapse. Finally, it was unknown if, and if so, what kind of treatment patients had received. Currently, there is no universal accepted treatment for PCC, so the mainstay treatments of PCC are symptom management, rehabilitation, reassurance and support, and therapies to reduce sympathetic hyperactivity (30). Received treatment might differentially affect recovery from PCC, as receiving an effective treatment enhances the chance of recovery and thus subsequently leads to an underestimation of recovery rates without treatment.

### Conclusion

Only one in ten of severely impaired and only one in three patients with milder PCC recovered in a nine-month period, urgently calling for therapeutic options for PCC patients. Recovery was less likely when patients already had worse general, physical, mental or social health, adding to the complexity of problems that patients with PCC face during their illness. Future research should focus on identifying changeable factors, including biomedical factors not assessed in this study, that assist recovery and improve quality of life of PCC patients. Furthermore, we recommend a clinical PCC definition that differentiates by severity, such as based on capacity of daily functioning.

## Acknowledgements

We gratefully acknowledge S Brinkhues, M Van Herck, K Konings, LCJ Steijvers, CPB Moonen, S Mujakovic, HLG ter Waarbeek, N Bouwmeester-Vincken, MA Spruit, and AW Vaes for their valuable contribution throughout the shaping and data collection of the PRIME post-COVID study.

## Author contributions

Conceptualization and methodology were performed by D.P., C.v.B., C.d.H., C.H., and N.D-M. Data collection and processing was done by D. P, C.v.B., and S.W. Data visualization, formal analysis and the original draft was written by D.P. Reviewing and editing were performed by C.v.B., S.W., C.d.H., N.D-M., and C.H. N.D-M. supervised the study and acquired funding.

