## supplementary tables 1 and 2 for "Recovery from post-COVID-19 condition and associated factors: the PRIME post-COVID study"

Supplementary information

### Supplementary table 1. Factors associated with changing PCC status from post-COVID-19 condition with no or slight problems with performing daily activities (PCC-affectedDA)

|  | **Patients with PCC-affectedDA at baseline (n=602)** | | | | | | | |
| --- | --- | --- | --- | --- | --- | --- | --- | --- |
|  | Recovered (n=222) | | | PCC-affectedDA (n=321) ref. | | PCC impaired DA (n=59) | | |
| **Individual level** | n | % | adOR [95% CI] | n | % | n | % | adOR [95% CI] |
| *Demographic factors* |  |  |  |  |  |  |  |  |
| Women | 122 | 55.0 | 0.86 [0.61-1.22] | 188 | 58.6 | 40 | 67.8 | 1.54 [0.84-2.82] |
| Age, mean(SD) | 54 (13) | |  | 55 (13) | | 55 (10) | |  |
| Born in NL, (yes) | 204 | 91.9 | 0.64 [0.32-1.28] | 304 | 94.7 | 57 | 96.6 | 1.63 [0.36-7.26] |
| Level of education |  |  |  |  |  |  |  |  |
| Practically trained | 122 | 55.0 | ref. | 199 | 62.0 | 41 | 69.5 | ref. |
| Theoretically trained | 100 | 45.0 | 1.34 [0.95-1.91] | 122 | 38.0 | 18 | 30.5 | 0.72 [0.39-1.33] |
| Paid job (yes) | 163 | 73.4 | 1.16 [0.75-1.81] | 243 | 75.7 | 44 | 74.6 | 1.03 [0.51-2.08] |
| *General (physical) health status* |  |  |  |  |  |  |  |  |
| Obesity (yes) | 63 | 28.4 | 0.95 [0.65-1.38] | 95 | 29.6 | 21 | 35.6 | 1.30 [0.72-2.34] |
| Diabetes (yes) | 15 | 6.8 | 0.89 [0.45-1.75] | 24 | 7.5 | 4 | 6.8 | 0.86 [0.28-2.62] |
| COPD (yes) | 6 | 2.7 | 0.61 [0.23-1.61] | 14 | 4.4 | 3 | 5.1 | 1.19 [0.33-4.30] |
| Asthma (yes) | 13 | 5.9 | 0.75 [0.37-1.50] | 24 | 7.5 | 5 | 8.5 | 1.11 [0.40-3.06] |
| Lung disease (yes) | 4 | 1.8 | 1.97 [0.43-9.01] | 3 | 0.9 | 0 | 0.0 | - |
| Heart disease (yes) | 7 | 3.2 | 0.63 [0.25-1.60] | 15 | 4.7 | 3 | 5.1 | 1.05 [0.29-3.79] |
| General health score, mean(SD) |  |  |  |  |  |  |  |  |
| Year before test | 83.7 (11.9) | | **0.98 [0.98-0.99]**** | 82.9 (15.6) | | 79.8 (16.6) | | 1.01 [1.00-1.03] |
| At baseline | 75.3 (15.8) | | **0.98 [0.97-0.98]**** | 74.4 (15.4) | | 67.9 (15.2) | | **1.02 [1.01-1.04]*** |
| Fatigue |  |  |  |  |  |  |  |  |
| Normal | 101 | 45.5 | ref. | 90 | 28.0 | 10 | 16.9 | ref. |
| Mild | 62 | 27.9 | **0.51 [0.33-0.78]*** | 105 | 32.7 | 14 | 23.7 | 1.19 [0.50-2.83] |
| Severe | 59 | 26.6 | **0.40 [0.26-0.61]**** | 126 | 39.3 | 35 | 53.9 | **2.68 [1.23-5.81]*** |
| Dyspnea |  |  |  |  |  |  |  |  |
| Normal to mild | 211 | 95.0 | ref. | 298 | 92.8 | 48 | 81.4 | ref. |
| Severe | 11 | 5.0 | 0.67 [0.32-1.42] | 23 | 7.2 | 11 | 18.6 | **2.92 [1.33-6.42]*** |
| *Symptoms at baseline* |  |  |  |  |  |  |  |  |
| Amnesia |  |  |  |  |  |  |  |  |
| No | 207 | 93.2 | ref. | 280 | 87.2 | 47 | 79.7 | ref. |
| Severity 1-4 | 8 | 3.6 | 0.73 [0.30-1.75] | 15 | 4.7 | 6 | 10.2 | 2.32 [0.85-6.31] |
| Severity ≥5-10 | 7 | 3.2 | **0.37 [0.16-0.86]*** | 26 | 8.1 | 6 | 10.2 | 1.41 [0.55-3.62] |
| Brain fog |  |  |  |  |  |  |  |  |
| No | 213 | 95.9 | ref. | 298 | 92.8 | 55 | 93.2 | ref. |
| Severity 1-4 | 3 | 1.4 | 0.46 [0.12-1.73] | 9 | 2.8 | 1 | 1.7 | 0.60 [0.07-4.85] |
| Severity ≥5-10 | 6 | 2.7 | 0.62 [0.23-1.65] | 14 | 4.4 | 3 | 5.1 | 1.24 [0.34-4.55] |
| Concentration difficulties |  |  |  |  |  |  |  |  |
| No | 182 | 82.0 | ref. | 230 | 71.7 | 39 | 66.1 | ref. |
| Severity 1-4 | 17 | 7.7 | 0.70 [0.37-1.30] | 31 | 9.7 | 4 | 6.8 | 0.78 [0.26-2.32] |
| Severity ≥5-10 | 23 | 10.4 | **0.48 [0.29-0.81]*** | 60 | 18.7 | 16 | 27.1 | 1.60 [0.83-3.07] |
| Confusion |  |  |  |  |  |  |  |  |
| No | 219 | 98.6 | ref. | 314 | 97.8 | 57 | 96.6 | - |
| Severity 1-4 | 2 | 0.9 | 0.98 [0.16-5.91] | 3 | 0.9 | 2 | 3.4 |  |
| Severity ≥5-10 | 1 | 0.5 | 0.36 [0.04-3.22] | 4 | 1.2 | 0 | 0.0 |  |
| Dizziness |  |  |  |  |  |  |  |  |
| No | 205 | 92.3 | ref. | 296 | 92.2 | 53 | 89.8 | ref. |
| Severity 1-4 | 10 | 4.5 | 1.34 [0.56-3.22] | 11 | 3.4 | 5 | 8.5 | 2.57 [0.86-7.71] |
| Severity ≥5-10 | 7 | 3.2 | 0.73 [0.29-1.86] | 14 | 4.4 | 1 | 1.7 | 0.39 [0.05-3.03] |
| Headache |  |  |  |  |  |  |  |  |
| No | 201 | 90.5 | ref. | 276 | 86.0 | 47 | 79.7 | ref. |
| Severity 1-4 | 7 | 3.2 | 0.58 [0.23-1.45] | 16 | 5.0 | 3 | 5.1 | 1.11 [0.31-3.99] |
| Severity ≥5-10 | 14 | 6.3 | 0.68 [0.35-1.32] | 29 | 9.0 | 9 | 15.3 | 1.91 [0.84-4.35] |
| Palpitations |  |  |  |  |  |  |  |  |
| No | 210 | 94.6 | ref. | 297 | 92.5 | 53 | 89.8 | ref. |
| Severity 1-4 | 8 | 3.6 | 0.94 [0.38-2.35] | 12 | 3.7 | 4 | 6.8 | 1.83 [0.56-5.96] |
| Severity ≥5-10 | 4 | 1.8 | 0.49 [0.15-1.53] | 12 | 3.7 | 2 | 3.4 | 0.96 [0.21-4.44] |
| Increased resting heart rate |  |  |  |  |  |  |  |  |
| No | 216 | 97.3 | ref. | 308 | 96.0 | 54 | 91.5 | ref. |
| Severity 1-4 | 2 | 0.9 | 0.35 [0.07-1.65] | 8 | 2.5 | 3 | 5.1 | 2.04 [0.52-8.00] |
| Severity ≥5-10 | 4 | 1.8 | 1.18 [0.31-4.46] | 5 | 1.6 | 2 | 3.4 | 2.28 [0.43-12.11] |
| Joint pain |  |  |  |  |  |  |  |  |
| No | 220 | 99.1 | - | 314 | 97.8 | 56 | 94.9 | - |
| Severity 1-4 | 1 | 0.5 |  | 0 | 0.0 | 2 | 3.4 |  |
| Severity ≥5-10 | 1 | 0.5 |  | 7 | 2.2 | 1 | 1.7 |  |
| Muscle pain or -weakness |  |  |  |  |  |  |  |  |
| No | 209 | 94.1 | ref. | 278 | 86.6 | 51 | 86.4 | ref. |
| Severity 1-4 | 6 | 2.7 | 0.79 [0.28-2.21] | 10 | 3.1 | 2 | 3.4 | 1.05 [0.22-4.96] |
| Severity ≥5-10 | 7 | 3.2 | **0.28 [0.12-0.64]*** | 33 | 10.3 | 6 | 10.2 | 0.97 [0.38-2.44] |
| Sleeping problems |  |  |  |  |  |  |  |  |
| No | 191 | 86.5 | ref. | 262 | 81.6 | 44 | 74.6 | ref. |
| Severity 1-4 | 9 | 4.1 | 0.97 [0.40-2.32] | 13 | 4.0 | 5 | 8.5 | 2.38 [0.80-7.09] |
| Severity ≥5-10 | 21 | 9.5 | 0.62 [0.36-1.08] | 46 | 14.3 | 10 | 16.9 | 1.31 [0.62-2.80] |
| Loss/change of smell/taste |  |  |  |  |  |  |  |  |
| No | 169 | 76.1 | ref. | 214 | 66.7 | 35 | 59.3 | ref. |
| Severity 1-4 | 40 | 18.0 | 0.75 [0.48-1.16] | 66 | 20.6 | 14 | 23.7 | 1.27 [0.64-2.51] |
| Severity ≥5-10 | 13 | 5.9 | **0.39 [0.20-0.75]*** | 41 | 12.8 | 10 | 16.9 | 1.46 [0.66-3.21] |
| *Mobility* |  |  |  |  |  |  |  |  |
| Time spend lying down (hours/day) | 1.5 (2.7) | | **0.94 [0.91-0.97]**** | 1.5 (2.6) | | 1.9 (2.4) | | 1.04 [0.94-1.15] |
| Problems with walking |  |  |  |  |  |  |  |  |
| No to slight | 203 | 91.4 | ref. | 288 | 89.7 | 45 | 76.3 | ref. |
| Moderate to(very)severe | 19 | 8.6 | 0.81 [0.44-1.48] | 33 | 10.3 | 14 | 23.7 | **2.71 [1.32-5.54]*** |
| *(severity of) acute COVID-19 illness* |  |  |  |  |  |  |  |  |
| Symptoms when tested, mean (SD) | 7.2 (5.1) | | **0.89 [0.88-0.91]**** | 8.3 (6.6) | | 9.6 (6.0) | | 1.03 [0.99-1.07] |
| Hospitalization/oxygen use at home or hospital (yes) | 8 | 3.6 | **0.35 [0.16-0.78]*** | 31 | 9.7 | 9 | 15.3 | 1.71 [0.78-3.87] |
| Months since test, mean (SD) |  |  |  |  |  |  |  |  |
| 3-6 | 22 | 9.9 | - | 19 | 5.9 | 8 | 13.6 | - |
| 6-12 | 152 | 68.5 |  | 234 | 72.9 | 36 | 61.0 |  |
| >12 | 48 | 21.6 |  | 68 | 21.2 | 15 | 25.4 |  |
| New COVID-19 illness between baseline and follow-up (yes) | 62 | 27.9 | - | 96 | 29.9 | 15 | 25.4 | - |
| *Vaccination status* |  |  |  |  |  |  |  |  |
| COVID-19 vaccine (yes) | 217 | 97.7 | 1.93 [0.67-5.58] | 308 | 96.0 | 58 | 98.3 | 2.56 [0.32-20.18] |
| New COVID-19 vaccine between baseline and follow-up (yes) | 1 | 0.5 | 0.73 [0.06-8.29] | 2 | 0.6 | 1 | 1.7 | 2.98 [0.26-34.04] |
| *Mental health status* |  |  |  |  |  |  |  |  |
| Depression (yes) | 64 | 28.8 | **0.55 [0.38-0.79]**** | 135 | 42.1 | 32 | 54.2 | 1.67 [0.95-2.96] |
| Loneliness |  |  |  |  |  |  |  |  |
| Not lonely | 141 | 63.5 | ref. | 198 | 61.7 | 31 | 52.5 | ref. |
| Moderately lonely | 65 | 29.3 | 0.98 [0.67-1.44] | 94 | 29.3 | 21 | 35.6 | 1.46 [0.79-2.68] |
| Severely lonely | 16 | 7.2 | 0.77 [0.41-1.48] | 29 | 9.0 | 7 | 11.9 | 1.51 [0.60-3.77] |
| Emotional loneliness (yes) | 48 | 21.6 | 0.97 [0.64-1.47] | 72 | 22.4 | 15 | 25.4 | 1.19 [0.63-2.27] |
| Social loneliness (yes) | 49 | 22.1 | 0.81 [0.54-1.22] | 83 | 25.9 | 18 | 30.5 | 1.26 [0.68-2.31] |
| Anxiety (yes) | 0 | 0.0 | - | 9 | 2.8 | 0 | 0.0 | - |
| *Lifestyle* |  |  |  |  |  |  |  |  |
| Smoking behaviour |  |  |  |  |  |  |  |  |
| Never | 114 | 51.4 | ref. | 161 | 50.2 | 31 | 52.5 | ref. |
| Former | 87 | 39.2 | **0.80 [0.66-0.96]*** | 140 | 43.6 | 23 | 39.0 | 0.82 [0.44-1.53] |
| Current | 21 | 9.5 | 0.79 [0.57-1.10] | 20 | 6.2 | 5 | 8.5 | 1.26 [0.44-3.65] |
| **Interpersonal level** |  |  |  |  |  |  |  |  |
| *Steady partner* |  |  |  |  |  |  |  |  |
| Having a relationship (yes) | 173 | 77.9 | **0.61 [0.39-0.95]*** | 274 | 85.4 | 52 | 88.1 | 1.28 [0.55-2.99] |
| *Home situation* |  |  |  |  |  |  |  |  |
| Living alone (yes) | 44 | 19.8 | 1.30 [0.83-2.02] | 52 | 16.2 | 6 | 10.2 | 0.59 [0.24-1.44] |
| Children (yes) | 77 | 34.7 | 1.17 [0.80-1.71] | 101 | 31.5 | 18 | 30.5 | 0.97 [0.52-1.83] |
| *Social network factors* |  |  |  |  |  |  |  |  |
| Size of social network, n(SD) | 10.8 (7.3) | | 1.0 [0.99-1.01] | 10.2 (7.1) | | 9.9 (5.4) | | 1.00 [0.95-1.04] |
| Support from social network, n(SD) |  |  |  |  |  |  |  |  |
| Number of members giving emotional support | 7.5 (6.3) | | 0.99 [0.98-1.00] | 7.6 (6.00 | | 6.9 (4.8) | | 0.98 [0.93-1.03] |
| Number of members giving practical support | 1.9 (2.1) | | **0.94 [0.91-0.98]**** | 2.1 (2.2) | | 1.9 (1.6) | | 0.95 [0.83-1.10] |
| Number of members giving informational support | 5.1 (4.6) | | 1.00 [0.98-1.02] | 5.1 (4.7) | | 4.3 (3.6) | | 0.96 [0.89-1.03] |
| Network diversity |  |  |  |  |  |  |  |  |
| Family and friends and acquaintances/caregivers | 143 | 64.4 | ref. | 181 | 56.4 | 40 | 67.8 | ref. |
| Only family, no friends/acquaintances/caregivers | 24 | 10.8 | 0.77 [0.44-1.34] | 39 | 12.1 | 6 | 10.2 | 0.69 [0.27-1.75] |
| Other | 55 | 24.8 | 0.69 [0.46-1.02] | 101 | 31.5 | 13 | 22.0 | 0.58 [0.30-1.13] |
| Network density |  |  |  |  |  |  |  |  |
| Best friends know family (yes) | 189 | 85.1 | 1.19 [0.74-1.92] | 267 | 83.2 | 52 | 88.1 | 1.56 [0.67-3.66] |
| **Societal level** |  |  |  |  |  |  |  |  |
| *Physical environment* |  |  |  |  |  |  |  |  |
| Urbanity of living area |  |  |  |  |  |  |  |  |
| (very) strongly urban | 100 | 45.0 | ref. | 109 | 34.0 | 17 | 28.8 | ref. |
| Moderately urban | 47 | 21.2 | 0.68 [0.43-1.06] | 76 | 23.7 | 14 | 23.7 | 1.15 [0.53-2.48] |
| Little urban | 48 | 21.6 | 0.67 [0.43-1.06] | 77 | 24.0 | 15 | 25.4 | 1.20 [0.56-2.57] |
| Rural | 27 | 12.2 | **0.49 [0.29-0.83]*** | 59 | 18.4 | 13 | 22.0 | 1.39 [0.63-3.07] |
| *p<0.05 **p<0.001 PCC-affectedDA=post-COVID-19 condition with no or slight problems with performing daily activities; PCC-impairedDA=post-COVID-19 condition with moderate to severe problems with performing daily activities. Adjusted for age (years), time since test (3-6, 6-12, >12 months), and having had a new COVID-19 infection between baseline and follow-up (yes/no). | | | | | | | | |

### Supplementary table 2. Factors associated with changing PCC status from post-COVID-19 condition with moderate to severe problems with performing daily activities (PCC-impairedDA)

|  | **Patients with PCC-impairedDA at baseline (n=277)** | | | | | | | |
| --- | --- | --- | --- | --- | --- | --- | --- | --- |
|  | Recovered (n=30) | | | PCC-affectedDA (n=88) | | | PCC-impairedDA (n=159) ref. | |
| **Individual level** | n | % | adOR [95% CI] | n | % | adOR [95% CI] | n | % |
| *Demographic factors* |  |  |  |  |  |  |  |  |
| Women | 18 | 60.0 | 0.72 [0.31-1.67] | 56 | 63.6 | 0.85 [0.48-1.49] | 106 | 66.7 |
| Age, mean(SD) | 56 (11) | |  | 56 (13) | |  | 57 (11) | |
| Born in NL, (yes) | 27 | 90.0 | 0.39 [0.09-1.77] | 86 | 97.7 | 1.66 [0.33-8.47] | 153 | 96.2 |
| Level of education |  |  |  |  |  |  |  |  |
| Practically trained | 18 | 60.0 | ref. | 55 | 62.5 | ref. | 110 | 69.2 |
| Theoretically trained | 12 | 40.0 | 1.39 [0.60-3.20] | 33 | 37.5 | 1.31 [0.75-2.31] | 49 | 30.8 |
| Paid job (yes) | 19 | 63.3 | 0.84 [0.32-2.18] | 47 | 53.4 | 1.45 [0.79-2.64] | 92 | 57.9 |
| *General (physical) health status* |  |  |  |  |  |  |  |  |
| Obesity (yes) | 9 | 30.0 | 0.72 [0.31-1.70] | 31 | 35.2 | 0.89 [0.52-1.54] | 61 | 38.4 |
| Diabetes (yes) | 2 | 6.7 | 0.60 [0.13-2.77] | 6 | 6.8 | 0.54 [0.21-1.43] | 19 | 11.9 |
| COPD (yes) | 3 | 10.0 | 1.44 [0.37-5.59] | 5 | 5.7 | 0.67 [0.23-1.94] | 14 | 8.8 |
| Asthma (yes) | 3 | 10.0 | 0.55 [0.15-1.97] | 11 | 12.5 | 0.73 [0.34-1.56] | 26 | 16.4 |
| Lung disease (yes) | 2 | 6.7 | 3.30 [0.49-22.41] | 0 | 0.0 | - | 3 | 1.9 |
| Heart disease (yes) | 0 | 0.0 | - | 10 | 11.4 | 1.90 [0.75-4.78] | 11 | 6.9 |
| General health score, mean(SD) |  |  |  |  |  |  |  |  |
| Year before test | 77.8 (17.5) | | 0.99 [0.97-1.01] | 75.5 (20.4) | | 1.00 [0.98-1.01] | 73.5 (22.4) | |
| At baseline | 53.5 (18.5) | | 0.98 [0.96-1.00] | 58.6 (17.5) | | **0.97 [0.95-0.98]**** | 48.2 (20.3) | |
| Fatigue |  |  |  |  |  |  |  |  |
| Normal | 3 | 10.0 | ref. | 6 | 6.8 | ref. | 1 | 0.6 |
| Mild | 5 | 16.7 | **0.07 [0.01-0.98]*** | 12 | 13.6 | **0.09 [0.01-0.81]*** | 19 | 11.9 |
| Severe | 22 | 73.3 | **0.03 [0.01-0.38]*** | 70 | 79.5 | **0.06 [0.01-0.53]*** | 139 | 87.4 |
| Dyspnea |  |  |  |  |  |  |  |  |
| Normal to mild | 26 | 86.7 | ref. | 56 | 63.6 | ref. | 90 | 56.6 |
| Severe | 4 | 13.3 | **0.19 [0.06-0.58]*** | 32 | 36.4 | 0.75 [0.43-1.28] | 69 | 43.4 |
| *Symptoms at baseline* |  |  |  |  |  |  |  |  |
| Amnesia |  |  |  |  |  |  |  |  |
| No | 28 | 93.3 | ref. | 60 | 68.2 | ref. | 93 | 58.5 |
| Severity 1-4 | 1 | 3.3 | 0.16 [0.02-1.30] | 9 | 10.2 | 0.78 [0.32-1.87] | 17 | 10.7 |
| Severity ≥5-10 | 1 | 3.3 | **0.05 [0.01-0.38]*** | 19 | 21.6 | 0.55 [0.29-1.05] | 49 | 30.8 |
| Brain fog |  |  |  |  |  |  |  |  |
| No | 26 | 86.7 | - | 73 | 83.0 | ref. | 109 | 68.6 |
| Severity 1-4 | 0 | 0.0 |  | 3 | 3.4 | 0.60 [0.15-2.42] | 7 | 4.4 |
| Severity ≥5-10 | 4 | 13.3 |  | 12 | 13.6 | **0.35 [0.17-0.74]*** | 43 | 27.0 |
| Concentration difficulties |  |  |  |  |  |  |  |  |
| No | 22 | 73.3 | ref. | 41 | 46.6 | ref. | 55 | 34.6 |
| Severity 1-4 | 4 | 13.3 | 0.59 [0.17-2.09] | 10 | 11.4 | 0.86 [0.34-2.17] | 14 | 8.8 |
| Severity ≥5-10 | 4 | 13.3 | **0.07 [0.02-0.24]**** | 37 | 42.0 | **0.48 [0.27-0.87]*** | 90 | 56.6 |
| Confusion |  |  |  |  |  |  |  |  |
| No | 29 | 96.7 | - | 79 | 89.8 | ref. | 141 | 88.7 |
| Severity 1-4 | 1 | 3.3 |  | 2 | 2.3 | 0.57 [0.11-2.92] | 6 | 3.8 |
| Severity ≥5-10 | 0 | 0.0 |  | 7 | 8.0 | 1.01 [0.38-2.69] | 12 | 7.5 |
| Dizziness |  |  |  |  |  |  |  |  |
| No | 27 | 90.0 | ref. | 69 | 78.4 | ref. | 119 | 74.8 |
| Severity 1-4 | 2 | 6.7 | 0.63 [0.13-2.97] | 10 | 11.4 | 1.20 [0.51-2.83] | 15 | 9.4 |
| Severity ≥5-10 | 1 | 3.3 | 0.18 [0.02-1.37] | 9 | 10.2 | 0.59 [0.26-1.35] | 25 | 15.7 |
| Headache |  |  |  |  |  |  |  |  |
| No | 25 | 83.3 | ref. | 67 | 76.1 | ref. | 109 | 68.6 |
| Severity 1-4 | 1 | 3.3 | 0.42 [0.05-3.40] | 7 | 8.0 | 0.89 [0.33-2.41] | 12 | 7.5 |
| Severity ≥5-10 | 4 | 13.3 | 0.49 [0.16-1.53] | 14 | 15.9 | 0.55 [0.27-1.12] | 38 | 23.9 |
| Palpitations |  |  |  |  |  |  |  |  |
| No | 30 | 100.0 | - | 75 | 85.2 | ref. | 133 | 83.6 |
| Severity 1-4 | 0 | 0.0 |  | 6 | 6.8 | 0.75 [0.28-2.04] | 14 | 8.8 |
| Severity ≥5-10 | 0 | 0.0 |  | 7 | 8.0 | 0.98 [0.37-2.62] | 12 | 7.5 |
| Increased resting heart rate |  |  |  |  |  |  |  |  |
| No | 26 | 86.7 | ref. | 76 | 86.4 | ref. | 125 | 78.6 |
| Severity 1-4 | 2 | 6.7 | 1.60 [0.31-8.34] | 4 | 4.5 | 0.81 [0.23-2.81] | 8 | 5.0 |
| Severity ≥5-10 | 2 | 6.7 | 0.37 [0.08-1.67] | 8 | 9.1 | 0.49 [0.21-1.15] | 26 | 16.4 |
| Joint pain |  |  |  |  |  |  |  |  |
| No | 28 | 93.3 | - | 78 | 88.6 | ref. | 143 | 89.9 |
| Severity 1-4 | 0 | 0.0 |  | 2 | 2.3 | 1.80 [0.24-13.43] | 2 | 1.3 |
| Severity ≥5-10 | 2 | 6.7 |  | 8 | 9.1 | 1.01 [0.40-2.53] | 14 | 8.8 |
| Muscle pain or -weakness |  |  |  |  |  |  |  |  |
| No | 25 | 83.3 | - | 58 | 65.9 | ref. | 102 | 64.2 |
| Severity 1-4 | 0 | 0.0 |  | 3 | 3.4 | 0.50 [0.13-1.92] | 10 | 6.3 |
| Severity ≥5-10 | 5 | 16.7 |  | 27 | 30.7 | 1.01 [0.57-1.81] | 47 | 29.6 |
| Sleeping problems |  |  |  |  |  |  |  |  |
| No | 24 | 80.0 | ref. | 61 | 69.3 | ref. | 96 | 60.4 |
| Severity 1-4 | 1 | 3.3 | 0.39 [0.05-3.19] | 3 | 3.4 | 0.43 [0.11-1.60] | 11 | 6.9 |
| Severity ≥5-10 | 5 | 16.7 | 0.37 [0.13-1.03] | 24 | 27.3 | 0.75 [0.42-1.34] | 52 | 32.7 |
| Loss/change of smell/taste |  |  |  |  |  |  |  |  |
| No | 22 | 73.3 | ref. | 51 | 58.0 | ref. | 98 | 61.6 |
| Severity 1-4 | 4 | 13.3 | 0.46 [0.14-1.48] | 22 | 25.0 | 1.23 [0.65-2.33] | 34 | 21.4 |
| Severity ≥5-10 | 4 | 13.3 | 0.49 [0.15-1.64] | 15 | 17.0 | 1.03 [0.50-2.26] | 27 | 17.0 |
| *Mobility* |  |  |  |  |  |  |  |  |
| Time spend lying down (hours/day) | 2.5 (2.4) | | 1.00 [0.88-1.14] | 2.5 (2.8) | | 1.00 [0.92-1.09] | 2.4 (3.2) | |
| Problems with walking |  |  |  |  |  |  |  |  |
| No to slight | 20 | 66.7 | ref. | 49 | 55.7 | ref. | 73 | 45.9 |
| Moderate to(very)severe | 10 | 33.3 | **0.40 [0.17-0.97]*** | 39 | 44.3 | 0.70 [0.40-1.21] | 86 | 54.1 |
| *(severity of) acute COVID-19 illness* |  |  |  |  |  |  |  |  |
| Symptoms when tested, mean (SD) | 7.6 (6.2) | | **0.92 [0.86-0.97]*** | 11.0 (8.3) | | 0.98 [0.95-1.01] | 12.7 (8.7) | |
| Hospitalization/oxygen use at home or hospital (yes) | 4 | 13.3 | 0.51 [0.16-1.58] | 11 | 12.5 | **0.43 [0.21-0.89]*** | 40 | 25.2 |
| Months since test, mean (SD) |  |  |  |  |  |  |  |  |
| 3-6 | 3 | 10.0 | - | 8 | 9.1 |  | 11 | 6.9 |
| 6-12 | 26 | 86.7 |  | 65 | 73.9 |  | 117 | 73.6 |
| >12 | 1 | 3.3 |  | 15 | 17.0 |  | 31 | 19.5 |
| New COVID-19 illness between baseline and follow-up (yes) | 4 | 13.3 | 0.99 [0.95-1.03] | 21 | 23.9 | 0.99 [0.97-1.02] | 35 | 22.0 |
| *Vaccination status* |  |  |  |  |  |  |  |  |
| COVID-19 vaccine (yes) | 26 | 86.7 | 0.37 [0.09-1.46] | 86 | 97.7 | 2.31 [0.46-11.70] | 152 | 95.6 |
| New COVID-19 vaccine between baseline and follow-up (yes) | 0 | 0.0 | - | 1 | 1.1 | 0.92 [0.08-10.44] | 2 | 1.3 |
| *Mental health status* |  |  |  |  |  |  |  |  |
| Depression (yes) | 20 | 66.7 | **0.29 [0.12-0.73]*** | 66 | 75.0 | **0.43 [0.22-0.85]*** | 137 | 86.2 |
| Loneliness |  |  |  |  |  |  |  |  |
| Not lonely | 15 | 50.0 | ref. | 54 | 61.4 | ref. | 74 | 46.5 |
| Moderately lonely | 9 | 30.0 | 0.70 [0.28-1.77] | 20 | 22.7 | **0.49 [0.27-0.92]*** | 55 | 34.6 |
| Severely lonely | 6 | 20.0 | 0.94 [0.33-1.70] | 15 | 15.9 | 0.62 [0.30-1.28] | 30 | 18.9 |
| Emotional loneliness (yes) | 10 | 33.3 | 0.76 [0.33-1.75] | 24 | 27.3 | **0.55 [0.31-0.98]*** | 63 | 39.6 |
| Social loneliness (yes) | 9 | 30.0 | 1.01 [0.42-2.40] | 20 | 22.7 | 0.71 [0.38-1.30] | 46 | 28.9 |
| Anxiety (yes) | 1 | 3.3 | 0.96 [0.11-8.49] | 2 | 2.3 | 0.61 [0.12-3.10] | 6 | 3.8 |
| *Lifestyle* |  |  |  |  |  |  |  |  |
| Smoking behaviour |  |  |  |  |  |  |  |  |
| Never | 16 | 53.3 | ref. | 47 | 53.4 | ref. | 81 | 50.9 |
| Former | 13 | 43.3 | 1.09 [0.46-2.56] | 35 | 39.8 | 0.99 [0.56-1.77] | 65 | 40.9 |
| Current | 1 | 3.3 | 0.41 [0.05-3.41] | 6 | 6.8 | 0.84 [0.30-2.39] | 13 | 8.2 |
| **Interpersonal level** |  |  |  |  |  |  |  |  |
| *Steady partner* |  |  |  |  |  |  |  |  |
| Having a relationship (yes) | 27 | 90.0 | 2.39 [0.65-8.72] | 71 | 80.7 | 0.99 [0.51-1.93] | 129 | 81.1 |
| *Home situation* |  |  |  |  |  |  |  |  |
| Living alone (yes) | 3 | 10.0 | 0.45 [0.12-1.65] | 17 | 19.3 | 1.09 [0.56-2.15] | 29 | 18.2 |
| Children (yes) | 10 | 33.3 | 0.86 [0.35-2.14] | 29 | 33.0 | 0.86 [0.47-1.56] | 54 | 34.0 |
| *Social network factors* |  |  |  |  |  |  |  |  |
| Size of social network, n(SD) | 11.4 (7.2) | | 0.98 [0.93-1.04] | 11.6 (7.4) | | 0.99 [0.96-1.02] | 12.4 (7.9) | |
| Support from social network, n(SD) |  |  |  |  |  |  |  |  |
| Number of members giving emotional support | 7.6 (5.4) | | 0.96 [0.90-1.03] | 8.5 (6.1) | | 0.99 [0.95-1.03] | 8.9 (6.7) | |
| Number of members giving practical support | 2.5 (1.3) | | 0.97 [0.84-1.12] | 3.1 (2.9) | | 1.03 [0.95-1.12] | 2.9 (3.5) | |
| Number of members giving informational support | 4.6 (3.7) | | 0.95 [0.87-1.04] | 5.7 (5.5) | | 1.00 [0.95-1.05] | 5.6 (5.3) | |
| Network diversity |  |  |  |  |  |  |  |  |
| Family and friends and acquaintances/caregivers | 20 | 66.7 | ref. | 53 | 60.2 | ref. | 93 | 58.5 |
| Only family, no friends/acquaintances/caregivers | 2 | 6.7 | 0.46 [0.10-2.19] | 6 | 6.8 | 0.57 [0.21-1.54] | 18 | 11.3 |
| Other | 8 | 26.7 | 0.85 [0.34-2.12] | 29 | 33.0 | 1.06 [0.60-1.88] | 48 | 30.2 |
| Network density |  |  |  | 6 |  |  |  |  |
| Best friends know family (yes) | 23 | 79.3 | 1.11 [0.41-2.98] | 73 | 83.9 | 1.31 [0.66-2.59] | 126 | 80.8 |
| **Societal level** |  |  |  |  |  |  |  |  |
| *Physical environment* |  |  |  |  |  |  |  |  |
| Urbanity of living area |  |  |  |  |  |  |  |  |
| (very) strongly urban | 8 | 26.7 | ref. | 25 | 28.4 | ref. | 60 | 37.7 |
| Moderately urban | 8 | 26.7 | 1.29 [0.44-3.82] | 24 | 27.3 | 1.49 [0.74-2.98] | 38 | 23.9 |
| Little urban | 5 | 16.7 | 1.17 [0.35-3.93] | 25 | 28.4 | 1.88 [0.93-3.80] | 33 | 20.8 |
| Rural | 9 | 30.0 | 2.28 [0.77-6.71] | 14 | 15.9 | 1.11 [0.50-2.49] | 28 | 17.6 |
| *p<0.05 **p<0.001 PCC-affectedDA=post-COVID-19 condition with no or slight problems with performing daily activities; PCC-impairedDA=post-COVID-19 condition with moderate to severe problems with performing daily activities. Adjusted for age (years), time since test (3-6, 6-12, >12 months), and having had a new COVID-19 infection between baseline and follow-up (yes/no). | | | | | | | | |
